# Structured psychiatric care and psychosocial support during placebo participation: association with violent and domestic-violence offending in the ReINVEST trial

**DOI:** 10.64898/2026.05.09.26352691

**Authors:** Emaediong I. Akpanekpo, Lee Knight, Mathew Gullotta, Peter W. Schofield, Tony Butler

**Author notes:** **Corresponding Author:** Emaediong I. Akpanekpo, UNSW Sydney, NSW 2052, Australia.

## Abstract

**Background:** Participants in the ReINVEST randomised placebo-controlled trial of sertraline, conducted among men with high trait impulsivity and histories of violent offending, received structured clinical contact throughout the trial, including psychiatric assessments, nursing consultations, crisis support, and referrals to mental health and external services. We estimated the effect of placebo trial participation, compared with non-participation after baseline and single-blind run-in, on violent and domestic-violence reoffending.

**Methods:** This prespecified secondary analysis included men from the ReINVEST trial pathway who completed baseline assessment and entered the single-blind run-in phase but did not proceed to randomisation, to inform the counterfactual. Violent and domestic-violence offences were identified from linked administrative records over 12- and 24-month follow-up periods. The adjusted difference in offending was estimated using two independent analytical approaches accounting for baseline differences. Additional analyses examined whether the effect varied by baseline clinical and criminal-history characteristics, whether pre-randomisation external referrals explained selection into placebo participation, and whether post-randomisation external referrals accounted for any part of the estimated effect.

**Results:** Placebo trial participation was associated with lower offending across both outcome domains and follow-up periods. Placebo-standardised mean count differences for violent offending were −0.19 (95% confidence interval [CI] −0.38, −0.04) at 12 months and −0.22 (95% CI −0.51, −0.05) at 24 months. Corresponding differences for domestic-violence offending were −0.37 (95% CI −0.81, −0.14) at 12 months and −0.49 (95% CI −0.92, −0.22) at 24 months. The association was more apparent among men with a documented psychiatric history and, for domestic-violence offending, among those with higher baseline anger, irritability and aggression. Pre-randomisation referrals did not explain selection into placebo participation or materially alter the estimates. Post-randomisation referrals were observed in both groups, remained more common in the placebo group, and did not account for the observed association.

**Conclusion:** Placebo participation in this trial involved sustained clinical contact and psychosocial support beyond exposure to inactive medication, and these non-pharmacological components may have contributed to lower reoffending. In placebo-controlled trials involving populations with high psychiatric morbidity and limited continuity of coordinated care, the clinical content of placebo participation should be explicitly characterised in trial design and interpretation.

**Trial Registration:** Australian New Zealand Clinical Trials Registry Identifier: ACTRN12613000442707

## INTRODUCTION

Interpersonal violence and domestic and family violence are major contributors to premature mortality, injury-related disability, psychological trauma, and societal cost (Karakurt, Smith and Whiting, 2014; Peterson *et al*., 2018; Vos *et al*., 2020; Zhou *et al*., 2024). Recidivism rates remain elevated in most countries, and the trajectory from initial conviction to reoffending is accelerated among individuals with complex clinical and social needs (Ogilvie *et al*., 2023; Yukhnenko *et al*., 2023; Akpanekpo *et al*., 2025; Syasyila *et al*., 2025). These characteristics are associated with reduced responsiveness to conventional interventions and with disproportionate contributions to population-level rates of violent crime (Falk *et al*., 2014; Moffitt, 2018). Despite sustained investment in correctional supervision and rehabilitation programmes, violent and domestic-violence reoffending have remained difficult to reduce (Gannon *et al*., 2019; Beaudry *et al*., 2021), generating interest in complementary approaches including pharmacological strategies aimed at the biological mechanisms underlying impulsive aggression (Huband *et al*., 2010; Fazel *et al*., 2014). These populations are also characterised by high levels of mental health morbidity and poor continuity of mental health care during the transition from custody to the community (Fazel *et al*., 2016; Browne *et al*., 2022). The intersection of psychiatric morbidity, fragmented service access, and elevated reoffending risk raises the possibility that structured clinical contact, provided within the context of a clinical trial, may represent a meaningful increment in psychiatric care for this population.

One candidate biological pathway involves serotonin regulation. Evidence from neurobiological, pharmacological, and genetic research has linked impaired serotonin signalling to increased impulsive and reactive aggression across a range of populations (Coccaro and Kavoussi, 1997; Caspi *et al*., 2002; Popova, 2006; Siever, 2008). These findings generated the hypothesis that selective serotonin reuptake inhibitors (SSRIs), which increase serotonin availability in the brain, might reduce impulsive aggression among individuals with chronically elevated impulsivity and histories of violent behaviour (Coccaro, 1989; Cherek *et al*., 2002; Coccaro, Lee and Kavoussi, 2009; Coccaro and Lee, 2019). However, existing trial evidence has largely come from small studies in psychiatric or forensic settings using laboratory measures or self-reported aggression as proxies for real-world behaviour, and it remains unclear whether effects on impulsivity translate into reductions in actual reoffending among community-based justice-involved populations. This evidence gap provided the rationale for the ReINVEST trial (Butler *et al*., 2021). ReINVEST was designed as a pharmacological efficacy trial, but its implementation involved substantial clinical support to facilitate retention and ensure duty of care (Morgan *et al*., 2025). Participants had sustained contact with psychiatrists, mental health nurses, and psychologists, and clinician-participant interactions extended beyond protocol to include case management, crisis support, service referral, and liaison with courts and other agencies (Butler *et al*., 2025). In the conventional logic of a placebo-controlled trial, the placebo arm is intended to estimate outcomes in the absence of pharmacological effect. However, in a trial where both arms received intensive and sustained clinical contact that extended beyond protocol, the placebo condition may not be equivalent to the absence of clinical input. Placebo participation plausibly represented exposure to a structured package of psychiatric monitoring, clinical support, referral, and advocacy that, for many participants, exceeded the level of coordinated mental health care they would otherwise have received. Post-trial linkage of trial records to the state reoffending database showed that offending rates were higher among men who did not proceed to randomisation than among those randomised to either placebo or sertraline (Butler *et al*., 2025). Although this observational finding was not protected by randomisation, the magnitude and consistency of the difference in regression-adjusted analyses raised the hypothesis that trial participation may have been associated with subsequent offending independently of pharmacological assignment.

This study was designed to address these gaps. The analysis was limited to placebo participants and men who completed the baseline assessment and entered the single-blind run-in phase but did not proceed to randomisation, thereby isolating the participation effect from pharmacological effects. The primary aim was to estimate the marginal difference in subsequent violent and domestic-violence reoffending associated with entry into the randomised trial structure under placebo, compared to not entering the randomised phase after baseline assessment and entry into the run-in phase. The second aim was to characterise the scope and frequency of clinician-delivered psychiatric and psychosocial supports recorded in the trial’s clinical management system. The third aim was to assess whether the estimated placebo participation effect varied across selected baseline clinical and criminal-history characteristics. The fourth aim was to examine whether external referrals recorded before randomisation explained selection into placebo participation, and whether external referrals recorded after randomisation accounted for any part of the estimated participation effect.

## METHODS

### Study design and data sources

This prespecified secondary analysis utilised data from the ReINVEST trial, a randomised, double-blind, placebo-controlled trial of sertraline in highly impulsive adult men with histories of violent offending. Trial records were linked to the New South Wales (NSW) Reoffending Database (ROD) to ascertain criminal justice outcomes over 12- and 24-month follow-up periods. The study used a causal inference approach based on potential outcomes (Hernan and Robins, 2024). The primary target of inference was the expected difference in offending that would have been observed among placebo participants had they not entered the randomised phase; that is, what would have happened to the same men under non-participation, with all other factors held constant. This quantity was chosen because the policy-relevant question concerns the impact of trial participation on those who actually enrolled, rather than an average across all men who might have been eligible. Valid estimation required three assumptions (Cole and Frangakis, 2009; Imbens and Rubin, 2015): (1) that participation status was independent of unmeasured confounders given the observed baseline covariates (conditional exchangeability); (2) that all participants had a non-zero probability of being in either group given their covariates (positivity); and (3) that the defined exposure corresponded to a well-specified intervention (consistency).

### Trial Registration and Ethics

The trial protocol has been published previously (Butler *et al*., 2021), and the primary results are reported elsewhere (Butler *et al*., 2025). The trial was registered with the Australian New Zealand Clinical Trials Registry (ACTRN12613000442707) and received ethical approval from the University of New South Wales (UNSW) Human Research Ethics Committee (HC11390, HC17771), the Aboriginal Health and Medical Research Council (822/11), Corrective Services NSW (09/26576), and NSW Justice Health and Forensic Mental Health Network (G8/14).

### Participants and trial procedures

Eligible participants were adult males aged 18 years or older who were identified through psychiatric assessment as having high impulsivity, defined by a Barratt Impulsiveness Scale total score of at least 70, and a documented history of at least two violent offences. Participants were recruited through referrals from courts, community corrections agencies, legal representatives, and self-referral. Exclusion criteria included inability to provide informed consent, major mental illness or clinically significant comorbidities, and current use of antipsychotic, anticonvulsant, or serotonergic medications.

Following screening and eligibility assessment, participants who met ongoing eligibility criteria entered a run-in phase during which all participants received open-label sertraline to assess tolerability and willingness to adhere to the follow-up schedule. Individuals who completed the run-in phase and met ongoing eligibility criteria were randomised to continue sertraline or transition to an identical placebo following down-titration. Participants attended protocol visits at baseline, at week two, and then every four weeks until approximately week 50. Clinical consultations were conducted by psychiatrists, mental health nurses, and psychologists throughout the randomised phase.

### Clinical team model and nature of trial participation

The clinical team consisted of mental health nurses and psychologists, supported by psychiatrists, who collectively provided psychiatric assessments and ongoing eligibility screenings, weekly telephone check-in calls, monthly in-person medication reviews, and access to a 24-hour crisis hotline. This structured clinical contact occurred on a regular, predictable schedule and represented sustained engagement with the health and support system throughout the duration of the trial. In addition to protocol-specified tasks, clinicians performed case management activities beyond the formal trial protocol, including advocacy and liaison with external services, referrals to community supports, crisis support extending to court and service attendances, as well as risk escalation and trauma-informed responses. This broader clinical-contact package is relevant to interpretation of the placebo participation effect because it constitutes a plausible non-pharmacologic component of placebo participation in this trial.

### Analytic groups and exposure definition

The exposure contrast was between participation in the placebo trial and non-participation during the randomised phase. The placebo group included all participants randomised to placebo as part of the double-blind phase. The counterfactual was informed by men who completed baseline assessment and entered the single-blind run-in phase but did not proceed to randomisation (hereafter, the comparison group). These men had entered the same pre-randomisation trial pathway but were not exposed to the subsequent structured clinical contact and repeated follow-up visits that characterised participation in the randomised phase.

### Outcomes and follow-up

All offending outcomes were ascertained by probabilistic linkage of trial records to the ROD, which captures proven criminal offences from court and incarceration records. Two offending outcome domains were assessed: violent offending and domestic-violence offending. Violent offences were classified using Australian and New Zealand Standard Offence Classification categories 01 to 06 (Australian Bureau of Statistics, 2011). Domestic-violence offences were identified using domestic-violence offence flags within the ROD. Outcomes were analysed over 12-month and 24-month follow-up horizons from the screening date. The outcome for each horizon was the number of proven offence events occurring during the relevant follow-up window, with multiple offences recorded on the same date treated as a single event and classified according to the most serious offence recorded on that date. Person-time at risk in the community during each window was incorporated as a log-transformed offset in all outcome models so that estimated differences reflect offending conditional on time at risk rather than crude differences in event accumulation (Osgood, 2000).

### Covariates

Adjustment covariates for the primary participation analyses were selected a priori as potential confounders of the participation-offending association. The baseline covariate set included Indigenous status, history of juvenile detention, any prior offending, prior violent offending, prior domestic-violence offending, psychiatric history, baseline depressive symptom severity, baseline anger and irritability, baseline psychological distress, and baseline self-reported physical and mental health functioning. Where analyses specifically examined the role of pre-randomisation external referral history, this set was supplemented by indicators of prior court assistance/support, prior medical services other than mental health services, prior mental health services, prior legal services, and any prior external referral.

### Statistical analysis

We estimated the placebo-standardised marginal difference in offending between placebo trial participation and non-participation. The primary analysis used common-support restriction with g-computation. A logistic regression model was fitted for the probability of belonging to the placebo group rather than the comparison group using the baseline covariate set. The analytic sample was then restricted to participants whose estimated propensity score lay within the range observed among placebo participants, to confine inference to the region of shared covariate support. Within this common-support sample, offending counts were modelled using Poisson regression with a participation indicator, the baseline covariates, and a log-transformed person-time offset. G-computation was used to generate predicted offending counts for each placebo participant under two scenarios: placebo participation as observed and counterfactual non-participation, with baseline covariates held at their observed values. The participation effect was estimated as the average difference between these two sets of predictions across placebo participants. A sensitivity analysis used entropy balancing to target the same estimand while retaining the full analytic sample (Snowden, Rose and Mortimer, 2011; Hainmueller, 2012; Zhao and Percival, 2017).

Effect-modification analyses were conducted at 12 months post-randomisation. Continuous modifiers included prior violent offence count, prior domestic-violence offence count, depressive symptom severity, anger and irritability, psychological distress, and self-reported mental health functioning. Each was measured at baseline and modelled using a natural cubic spline, with interaction terms used to assess whether the participation effect varied across the modifier distribution. Participation effects were estimated at the 25th, 50th, and 75th percentiles of each continuous modifier, and the higher-versus-lower contrast was defined as the difference between the effects estimated at the 75th and 25th percentiles. Psychiatric history was examined separately as a binary modifier. Joint Wald tests were used for interaction terms, and percentile confidence intervals were obtained by nonparametric bootstrap.

External referral activity was compared between placebo participants and non-randomised participants, with referral categories treated as non-mutually exclusive. Referrals recorded before randomisation were examined as potential confounders by: (1) modelling placebo participation as a function of the pre-randomisation referral variables and (2) repeating the main participation analyses after adding these measures to the adjustment set. Referrals recorded after randomisation were examined separately as candidate intermediate variables that might account for part of the association between placebo participation and later offending. Indirect, direct, and total components were estimated using single-mediator interventional analyses under the same common-support framework as the main analyses.

Uncertainty was quantified using nonparametric bootstrap resampling at the participant level with replacement. In each bootstrap replicate, the full estimation procedure was repeated from propensity score estimation through outcome modelling and g-computation. Percentile confidence intervals were derived from the empirical bootstrap distribution. All analyses were conducted in Python using standard scientific computing libraries.

## RESULTS

### Analytic sample and baseline balance

The analytic sample comprised 475 eligible men, including 309 in the placebo group and 166 in the comparison group (Figure 1). Restriction to the common-support region retained 468 participants (all 309 placebo and 159 of 166 comparison-group participants). Entropy balancing reduced measured baseline differences to negligible levels in both the whole sample and the common-support sample (Table 1).

**Table 1.** Baseline characteristics by analytic group, whole sample and common-support sample.

| Variable |  | Whole Sample (N = 475) |  |  |  | Common Support (N = 468) |  |  |  |
| --- | --- | --- | --- | --- | --- | --- | --- | --- | --- |
|  |  | Placebo (n = 309) | Entered run-in, not randomised (n = 166) | SMD (pre) | SMD (EB) | Placebo (n = 309) | Entered run-in, not randomised (n = 159) | SMD (pre) | SMD (EB) |
| **Sociodemographic** |  |  |  |  |  |  |  |  |  |
| Age (years) | Mean (SD) | 32.49 (9.95) | 31.05 (9.82) | 0.15 | 0.00 | 32.49 (9.95) | 31.26 (9.83) | 0.12 | 0.00 |
| Indigenous | N (%) | 86 (27.8%) | 53 (32.3%) | −0.10 | −0.01 | 86 (27.8%) | 48 (30.6%) | −0.06 | −0.01 |
| **Criminal History** |  |  |  |  |  |  |  |  |  |
| Prior juvenile detention | N (%) | 67 (21.9%) | 57 (35.0%) | −0.29 | −0.01 | 67 (21.9%) | 51 (32.7%) | −0.24 | −0.01 |
| Any prior offence | Mean (SD) | 10.83 (8.42) | 14.83 (12.13) | −0.38 | 0.00 | 10.83 (8.42) | 13.45 (10.18) | −0.28 | 0.00 |
| Prior violent offence | Mean (SD) | 2.52 (2.24) | 2.56 (2.07) | −0.02 | 0.00 | 2.52 (2.24) | 2.55 (2.08) | −0.01 | 0.00 |
| Prior DV offence | Mean (SD) | 2.81 (3.03) | 2.46 (2.68) | 0.12 | 0.00 | 2.81 (3.03) | 2.47 (2.66) | 0.12 | 0.00 |
| **Baseline Clinical Measures** |  |  |  |  |  |  |  |  |  |
| Psychiatric history | N (%) | 246 (80.1%) | 134 (80.7%) | −0.02 | 0.01 | 246 (80.1%) | 127 (79.9%) | 0.01 | 0.01 |
| BDI score | Mean (SD) | 11.20 (8.57) | 10.85 (8.24) | 0.04 | 0.02 | 11.20 (8.57) | 10.97 (8.19) | 0.03 | 0.02 |
| AIAQ total | Mean (SD) | 71.17 (23.09) | 74.91 (22.39) | −0.16 | 0.00 | 71.17 (23.09) | 74.51 (22.52) | −0.15 | 0.00 |
| K10 score | Mean (SD) | 15.12 (8.99) | 15.08 (7.99) | 0.01 | 0.01 | 15.12 (8.99) | 15.34 (7.97) | −0.03 | 0.01 |
| SF-12 PCS | Mean (SD) | 53.15 (7.18) | 53.28 (7.88) | −0.02 | −0.04 | 53.15 (7.18) | 53.16 (7.99) | 0.00 | −0.04 |
| SF-12 MCS | Mean (SD) | 40.64 (12.40) | 39.61 (12.04) | 0.08 | −0.02 | 40.64 (12.40) | 39.49 (12.10) | 0.09 | −0.02 |
| N (%) for binary variables; Mean (SD) for continuous variables. SMD (pre): unadjusted standardised mean difference; SMD (EB): post-weighting SMD after entropy balancing. BDI = Beck Depression Inventory; AIAQ = Anger, Irritability and Aggression Questionnaire; K10 = Kessler Psychological Distress Scale; SF-12 PCS/MCS = Short Form-12 Physical/Mental Component Scores; DV = domestic violence; ESS = effective sample size. Common support defined by minimum and maximum propensity scores among placebo participants. Entropy-balancing ESS = 129.3 (whole sample), 130.2 (common support); maximum weight = 4.63 and 4.48, respectively. |  |  |  |  |  |  |  |  |  |

**Figure 1.**
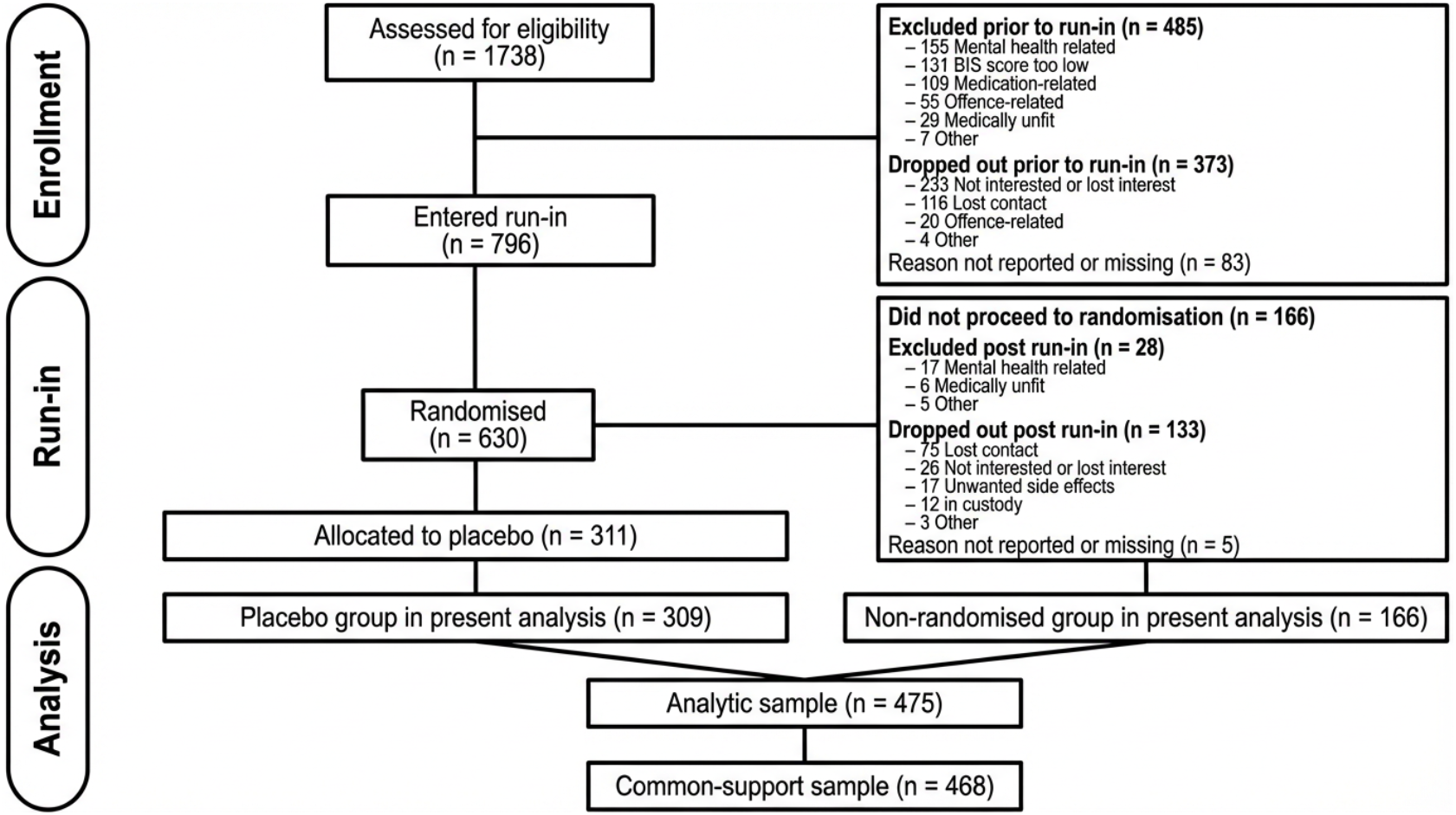
Flowchart of the study population.

### Association of placebo trial participation with reoffending

Placebo trial participation was associated with fewer violent and domestic-violence offences than non-participation under both estimators (Table 2). For violent offending, the placebo-standardised mean count difference was −0.19 (95% confidence interval [CI] −0.38 to −0.04) at 12 months and −0.22 (−0.51 to −0.05) at 24 months under common-support restriction. Estimates under entropy balancing were closely aligned. Differences for domestic-violence offending were larger in absolute magnitude: −0.37 (95% CI −0.81 to −0.14) at 12 months and −0.49 (−0.92 to −0.22) at 24 months under common support (Table 2).

**Table 2.** Placebo-standardised mean count differences for violent and domestic-violence reoffending (N = 468)

| Outcome | Follow-up period | Common-Support Effect (95% CI) | Entropy-Balanced Effect (95% CI) |
| --- | --- | --- | --- |
| Violent offending | 12 months | -0.19 (-0.38, -0.04) | -0.20 (-0.40, -0.02) |
|  | 24 months | -0.22 (-0.51, -0.05) | -0.23 (-0.50, -0.02) |
| DV offending | 12 months | -0.37 (-0.81, -0.14) | -0.37 (-0.70, -0.09) |
|  | 24 months | -0.49 (-0.92, -0.22) | -0.51 (-0.90, -0.17) |

### Effect-modification analyses

At 12 months, the estimated participation effect was larger among men with a documented psychiatric history than among those without, for both violent offending (−0.29 [95% CI −0.53 to −0.03] vs −0.13 [−0.56 to 0.14]) and domestic-violence offending (−0.79 [−1.39 to −0.15] vs −0.08 [−0.73 to 0.16]). For domestic-violence offending, the effect was also larger at the 75th percentile of baseline anger, irritability, and aggression (−0.66 [−1.19 to −0.12]) than at the 25th percentile (−0.34 [−0.99 to 0.08]). The higher-versus-lower contrasts were imprecise in all cases, and the interaction tests did not provide clear evidence of heterogeneity (Table 3).

**Table 3.** Placebo participation effects on 12-month offending outcomes by baseline characteristic level.

| Outcome | Modifier | Level | Common-Support Effect<br>(95% CI) | High vs. Low Contrast<br>(95% CI) | Interaction p-value |
| --- | --- | --- | --- | --- | --- |
| Violent offending | Psychiatric history | No history | −0.13 (−0.56, 0.14) | Reference | 0.400 |
|  |  | Any history | −0.29 (−0.53, −0.03) | −0.16 (−0.51, 0.32) |  |
| Domestic-violence<br>offending | Anger, Irritability and<br>Aggression Score | Low (P25) | −0.34 (−0.99, 0.08) | Reference | 1.000 |
|  |  | High (P75) | −0.66 (−1.19, −0.12) | −0.32 (−1.08, 0.43) |  |
|  | Psychiatric history | No history | −0.08 (−0.73, 0.16) | Reference | 0.082 |
|  |  | Any history | −0.79 (−1.39, −0.15) | −0.71 (−1.33, 0.30) |  |
| 12-month placebo-standardised mean count differences (common-support g-computation). Negative values indicate fewer offences under placebo. Continuous modifiers evaluated at the 25th and 75th percentiles using natural cubic splines. p-values are joint Wald tests for modifier-by-participation interaction. |  |  |  |  |  |

### Referrals to external services

Among participants with structured clinician-contact data, 179 placebo participants generated 524 referral events and 81 non-randomised participants generated 180 referral events (Table 4). Court assistance or support was the most frequent category in both groups, accounting for 53.6% of referral events in the placebo group and 59.4% in the non-randomised group. Medical services accounted for 21.4% and 19.4%, mental health services for 13.0% and 13.3%, and legal services for 7.4% and 4.4%, respectively (Table 4).

**Table 4.** Referrals to external services recorded by the ReINVEST clinical team.

| Referral category | Placebo | Placebo | Entered run-in, not randomised | Entered run-in, not randomised |
| --- | --- | --- | --- | --- |
|  | Events (n=524) | Participants (n=179) | Events (n=180) | Participants (n=81) |
| Court assistance/support | 281 (53.6%) | 137 (76.5%) | 107 (59.4%) | 61 (75.3%) |
| Medical services | 112 (21.4%) | 58 (32.4%) | 35 (19.4%) | 21 (25.9%) |
| Mental health services | 68 (13.0%) | 37 (20.7%) | 24 (13.3%) | 20 (24.7%) |
| Legal services | 39 (7.4%) | 24 (13.4%) | 8 (4.4%) | 7 (8.6%) |
| Police | 5 (1.0%) | 5 (2.8%) | 2 (1.1%) | 2 (2.5%) |
| Substance abuse/addiction | 11 (2.1%) | 5 (2.8%) | 0 (0%) | 0 (0%) |
| Charity organisations | 3 (0.6%) | 1 (0.6%) | 1 (0.6%) | 1 (1.2%) |
| Local community services | 1 (0.2%) | 1 (0.6%) | 2 (1.1%) | 2 (2.5%) |
| Disability services | 1 (0.2%) | 1 (0.6%) | 0 (0%) | 0 (0%) |
| Dept. Communities & Justice | 2 (0.4%) | 1 (0.6%) | 0 (0%) | 0 (0%) |
| Employment assistance | 0 (0%) | 0 (0%) | 1 (0.6%) | 1 (1.2%) |
| Victim support services | 1 (0.2%) | 1 (0.6%) | 0 (0%) | 0 (0%) |

When referrals were examined by timing relative to randomisation, external referrals before randomisation were more common in the comparison group across the main service categories (Supplementary Table 1). After randomisation, referrals were recorded in both groups but were more common in placebo participants: court assistance or support for 35.0% versus 16.9%, medical services for 15.2% versus 6.0%, mental health services for 8.1% versus 3.6%, and legal services for 5.5% versus 1.8% (Supplementary Table 1).

Pre-randomisation external referrals were examined as potential confounders of the participation-offending association. Prior court assistance or support was associated with a lower likelihood of later placebo participation (risk ratio 0.73, 95% CI 0.59 to 0.91), as was any prior external referral (0.73, 0.60 to 0.89) (Supplementary Table 2). However, additional adjustment for these variables produced little change in the estimated participation effects (Supplementary Table 3).

Post-randomisation external referrals were examined as candidate intermediate variables. Indirect components were small and their confidence intervals included zero across all referral categories and both estimators. For domestic-violence offending at 12 months, the total effect was −0.41 (95% CI −0.70 to −0.12) under common support with court assistance as the intermediate variable, and the corresponding indirect component was −0.00 (−0.04 to 0.04) (Supplementary Table 6). Results were similar across other referral categories and follow-up periods (Supplementary Tables 4-7). Post-randomisation external referrals did not account for the observed association.

## DISCUSSION

In this prespecified secondary analysis of the ReINVEST clinical trial, placebo participation was associated with fewer subsequent violent and domestic-violence offences than the counterfactual of non-participation. Estimated mean count differences were negative at 12 and 24 months for both outcome domains, with the largest absolute differences observed for domestic-violence offending. The referral profile included mental health services, medical care, and court assistance, consistent with a level of psychiatric care and psychosocial support that extended beyond the procedural requirements of a pharmacological trial. The estimates represent the average difference in offending associated with the full package of placebo participation, weighted to reflect the baseline characteristics of men who were actually randomised to placebo. Additional analyses of external referral activity did not materially alter this interpretation. Adjustment for referrals recorded before randomisation produced little change in the estimated participation effects, and referrals recorded after randomisation did not account for the association. These findings suggest that the observed association was not readily explained by differences in external service engagement alone and support interpretation of placebo participation as a broader clinical and psychosocial exposure.

The estimated participation effect was more clearly apparent among men with a documented psychiatric history and, for domestic-violence offending, at higher levels of baseline anger and irritability. These analyses were exploratory, the interaction tests did not provide clear evidence of heterogeneity, and the higher-versus-lower comparisons were imprecise. Even so, the direction of the findings is clinically coherent. In forensic psychiatric populations, anger-related constructs predict aggressive behaviour, and anger-control interventions are recognized treatment targets in the management of aggression, including in forensic psychiatric settings (Saini, 2009; Trestman, 2017). Anger and hostility have also been identified as relevant psychological factors in intimate partner violence perpetration (Birkley and Eckhardt, 2015). Psychiatric trial literature suggests that placebo-arm improvement is shaped in part by treatment context, including expectancy, therapeutic alliance, and other trial features such as visit structure and clinician contact (Walach *et al*., 2005; Leuchter *et al*., 2014; Wampold, 2018). It is plausible that men with psychiatric history and elevated anger-related symptoms were more likely to experience the trial’s structured monitoring, crisis support, and referral processes as a clinically meaningful increment in care. The present design cannot determine whether this trend reflects greater responsiveness to structured clinical contact, greater unmet need at baseline, or residual differences not fully captured by measured covariates, and these findings should therefore be interpreted as hypothesis-generating rather than confirmatory.

The present findings are consistent with a broader literature documenting that structured, intensive contact between justice-involved individuals and a consistent care or support team may itself be associated with lower reoffending trajectories (Kennealy *et al*., 2012; Trotter, 2013). Conceptually, this aligns with the common factors model of therapeutic change, which posits that relational and contextual elements of clinical encounters, including the therapeutic alliance, empathic attention, and the expectation of benefit, account for a substantial portion of improvement across therapeutic modalities, independent of the specific treatment delivered (Frank and Frank, 1991; Wampold, 2015). Durable reductions in violent and domestic-violence recidivism have proved difficult to achieve across evaluated programmes. Meta-analytic evidence from randomised trials of psychological interventions for incarcerated populations has shown modest aggregate effects that attenuate when small-study bias is accounted for (Beaudry *et al*., 2021), meta-analyses of batterer intervention programmes have reported weak effects on proven reoffending (Babcock, Green and Robie, 2004; Feder and Wilson, 2005), and a more recent meta-analysis of specialized offence-focused programmes estimated a 36% relative reduction in domestic-violence recidivism, though the evidence base draws largely on non-experimental designs (Gannon *et al*., 2019). In the present study, effect sizes for domestic-violence offending were similar across follow-up periods. However, these findings rely on the assumption that measured baseline characteristics sufficiently accounted for all systematic differences between groups, an assumption that cannot be verified.

These findings also have implications for the ethics of placebo-controlled trials in justice populations. A longstanding concern is that placebo assignment may deprive participants of active clinical benefit, an objection grounded in the principle of clinical equipoise (Freedman, 1987; Miller and Brody, 2002). In the ReINVEST context, this concern was mitigated by two features: no pharmacotherapy for impulsive aggression had established efficacy against violent reoffending at the time of trial design (Butler *et al*., 2021), satisfying the condition of genuine clinical uncertainty, and, as the present results suggest, placebo assignment was not equivalent to the absence of clinical input. Participants randomised to placebo received psychiatric assessment, clinical monitoring, crisis support, and referral to external services. The ethical implication is that placebo conditions in justice-involved trials may embed a more sustained clinical environment than is often acknowledged, and that differences in the continuity and intensity of that environment may matter even when participants who dropped out are not wholly unsupported. Ethical evaluation of placebo-controlled trials in these populations should consider the broader clinical content of both arms rather than focusing exclusively on whether an active drug is withheld.

### Strengths

This analysis has several methodological strengths. The counterfactual was informed by men drawn from the same trial pathway who completed the baseline assessment and entered the single-blind run-in phase but did not proceed to randomisation, providing a closer comparison than an external control group. Offending outcomes were ascertained from linked administrative records rather than self-report, reducing the risk of differential recall and social desirability bias. The estimand was prespecified and defined with respect to the baseline characteristics of men who entered the placebo arm. Common-support restriction limited comparisons to participants with overlapping covariate profiles, thereby reducing reliance on extrapolation, and the similarity of estimates under common-support restriction and entropy balancing indicated that the findings were not dependent on a single adjustment strategy. The external referral analyses further strengthened interpretation by addressing whether differences in earlier service engagement explained entry into placebo participation and by showing that the comparison group was not wholly unsupported after randomisation. That adjustment for pre-randomisation external referrals did not materially change the estimates, and that some post-randomisation external referral activity was also observed in the comparison group, makes the observed association more conservative than a comparison between a supported group and an unsupported one.

### Limitations

The principal limitation is that the contrast between placebo participation and non-participation was not protected by randomisation. Residual confounding by unmeasured factors, including motivation, accommodation stability, social support, and characteristics associated with attrition before randomisation, remains possible. Because the estimand was standardised to the placebo-group covariate distribution, the findings apply most directly to men with baseline characteristics similar to those who entered the placebo arm. The referral analyses were also limited by measurement issues. External referral categories captured only services outside the trial, were not mutually exclusive, and were sparse in some categories. They did not capture the structured psychiatric contact delivered directly by the trial team, which was the core component of the participation exposure. Accordingly, these analyses could narrow one measurable pathway but could neither identify the mechanism nor exclude pathways operating through the internal clinical structure of trial participation. Finally, the effect-modification analyses were exploratory, limited to the 12-month horizon, and not powered as primary tests of heterogeneity. Although continuous modifiers were modeled flexibly, the resulting estimates remained imprecise and should be interpreted cautiously.

## CONCLUSION

In the ReINVEST clinical trial, placebo participation was associated with lower subsequent violent and domestic-violence offending than non-participation. The association was evident at both 12 and 24 months, was not explained by differences in external referral activity before randomisation, and was not accounted for by external referrals recorded after randomisation. Although these analyses do not identify the mechanism, they are consistent with the view that the structured psychiatric contact and psychosocial support embedded in placebo participation constituted a clinically meaningful part of the trial condition. These findings underscore the importance of characterising the content of placebo participation when designing, reporting, and interpreting placebo-controlled trials in justice-involved populations.

## STATEMENTS AND DECLARATIONS

### Ethics Approval

All procedures performed in this study involving human participants were in accordance with the ethical standards of the institutional and national research committees and with the 1964 Declaration of Helsinki and its later amendments. Ethics approval was obtained from the University of New South Wales Human Research Ethics Committee (HC11390; HC17771), the Aboriginal Health and Medical Research Council Human Research Ethics Committee (822/11), the Corrective Services NSW Human Research Ethics Committee (09/26576), and the NSW Justice Health and Forensic Mental Health Network Human Research Ethics Committee (G8/14).

### Consent to Participate

Informed consent was obtained from all individual participants included in the study.

### Consent to Publish

Not applicable.

### Data Availability

The individual participant data generated and analysed during the current study are not publicly available because of specifications in participant consent forms regarding the sensitive nature of the data. De-identified data may be requested from the corresponding author, subject to relevant approvals.

## Acknowledgements

We thank the trial participants and the ReINVEST study team. We acknowledge the NSW Bureau of Crime Statistics and Research (BOCSAR) for conducting linkage to the Reoffending Database.

