## Supplementary Material for "Structured psychiatric care and psychosocial support during placebo participation: association with violent and domestic-violence offending in the ReINVEST trial"

**Supplementary Table 1. Participant-level external referrals in placebo and non-randomised groups before and after randomisation/index date**

| Referral category | Placebo total n (%) | Placebo before randomization n (%) | Placebo after randomization n (%) | Entered run-in, not randomised total n (%) | Entered run-in, not randomized, before index date n (%) | Entered run-in, not randomized, after index date n (%) |
|---|---|---|---|---|---|---|
| Court assistance/support | 134 (43.4) | 39 (12.6) | 108 (35.0) | 57 (34.3) | 40 (24.1) | 28 (16.9) |
| Medical services (non-mental health) | 53 (17.2) | 10 (3.2) | 47 (15.2) | 20 (12.0) | 12 (7.2) | 10 (6.0) |
| Mental health services | 30 (9.7) | 6 (1.9) | 25 (8.1) | 15 (9.0) | 9 (5.4) | 6 (3.6) |
| Legal services | 22 (7.1) | 5 (1.6) | 17 (5.5) | 7 (4.2) | 4 (2.4) | 3 (1.8) |
| Police | 5 (1.6) | 2 (0.6) | 3 (1.0) | 2 (1.2) | 1 (0.6) | 1 (0.6) |
| Substance abuse/addiction programs | 5 (1.6) | 2 (0.6) | 4 (1.3) | 0 (0.0) | 0 (0.0) | 0 (0.0) |
| Charity organisations | 1 (0.3) | 0 (0.0) | 1 (0.3) | 1 (0.6) | 0 (0.0) | 1 (0.6) |
| Department of Communities & Justice | 1 (0.3) | 1 (0.3) | 0 (0.0) | 0 (0.0) | 0 (0.0) | 0 (0.0) |
| Disability services | 1 (0.3) | 0 (0.0) | 1 (0.3) | 0 (0.0) | 0 (0.0) | 0 (0.0) |
| Local community organisations/services | 1 (0.3) | 0 (0.0) | 1 (0.3) | 2 (1.2) | 0 (0.0) | 2 (1.2) |
| Victim support services | 1 (0.3) | 0 (0.0) | 1 (0.3) | 0 (0.0) | 0 (0.0) | 0 (0.0) |
| Employment assistance | 0 (0.0) | 0 (0.0) | 0 (0.0) | 1 (0.6) | 1 (0.6) | 0 (0.0) |
| Aboriginal health/support services | 0 (0.0) | 0 (0.0) | 0 (0.0) | 0 (0.0) | 0 (0.0) | 0 (0.0) |
| Emergency services (non-medical) | 0 (0.0) | 0 (0.0) | 0 (0.0) | 0 (0.0) | 0 (0.0) | 0 (0.0) |
| Housing support | 0 (0.0) | 0 (0.0) | 0 (0.0) | 0 (0.0) | 0 (0.0) | 0 (0.0) |

The index date was defined as the screening date plus four weeks.

**Supplementary Table 2. Prior external referrals and later placebo participation**

| Prior referral variable | Risk ratio (95% CI) | P-value |
|---|---|---|
| Court assistance/support | 0.73 [0.59, 0.91] | 0.004 |
| Medical services (non-mental health) | 0.70 [0.45, 1.09] | 0.113 |
| Mental health services | 0.66 [0.37, 1.19] | 0.171 |
| Legal services | 0.81 [0.45, 1.46] | 0.488 |
| Any external referral | 0.73 [0.60, 0.89] | 0.002 |

Risk ratios are from models predicting later placebo participation from pre-randomisation referral variables. Risk ratios below 1.0 indicate lower likelihood of later placebo participation among those with the corresponding prior referral.

**Supplementary Table 3. Association between placebo participation and later offending before and after adjustment for prior external referrals**

| Outcome | Common support<br>Estimate (95% CI) | Entropy balanced<br>Estimate (95% CI) |
|---|---|---|
| Violent offending, 12 months | -0.20 [-0.43, -0.04] | -0.25 [-0.59, -0.02] |
| Violent offending, 24 months | -0.20 [-0.48, 0.01] | -0.26 [-0.65, 0.02] |
| Domestic-violence offending, 12 months | -0.41 [-0.67, -0.11] | -0.37 [-0.72, -0.07] |
| Domestic-violence offending, 24 months | -0.49 [-0.77, -0.13] | -0.45 [-0.81, -0.13] |

Estimates are standardised mean count differences. Negative values indicate fewer offending events among placebo participants. Prior-adjusted models added pre-randomisation external referral variables to the adjustment set.

**Supplementary Table 4. Interventional single-mediator analyses of post-randomisation external referrals for violent offending at 12 months**

| Mediator | Method | Indirect effect (95% CI) | Direct effect (95% CI) | Total effect (95% CI) |
|---|---|---|---|---|
| Court assistance/support | Common support | 0.02 [-0.01, 0.05] | -0.22 [-0.42, -0.09] | -0.20 [-0.42, -0.08] |
| Court assistance/support | Entropy balanced | 0.02 [-0.01, 0.05] | -0.27 [-0.53, -0.10] | -0.26 [-0.51, -0.07] |
| Medical services (non-mental health) | Common support | 0.00 [-0.01, 0.02] | -0.20 [-0.41, -0.07] | -0.20 [-0.41, -0.07] |
| Medical services (non-mental health) | Entropy balanced | 0.00 [-0.02, 0.02] | -0.25 [-0.51, -0.07] | -0.25 [-0.51, -0.07] |
| Mental health services | Common support | 0.00 [-0.01, 0.02] | -0.20 [-0.41, -0.07] | -0.20 [-0.41, -0.07] |
| Mental health services | Entropy balanced | 0.00 [-0.01, 0.02] | -0.25 [-0.52, -0.07] | -0.25 [-0.51, -0.06] |
| Legal services | Common support | 0.01 [-0.00, 0.03] | -0.21 [-0.41, -0.08] | -0.20 [-0.41, -0.07] |
| Legal services | Entropy balanced | 0.01 [-0.00, 0.04] | -0.26 [-0.53, -0.08] | -0.25 [-0.51, -0.07] |
| Event count | Common support | 0.03 [0.01, 0.05] | -0.23 [-0.43, -0.10] | -0.20 [-0.42, -0.07] |
| Event count | Entropy balanced | 0.03 [0.00, 0.05] | -0.29 [-0.54, -0.11] | -0.26 [-0.51, -0.08] |
| Distinct service categories | Common support | 0.02 [-0.00, 0.06] | -0.22 [-0.42, -0.09] | -0.20 [-0.41, -0.06] |
| Distinct service categories | Entropy balanced | 0.02 [-0.01, 0.06] | -0.27 [-0.52, -0.09] | -0.25 [-0.51, -0.06] |

Estimates are standardised mean count differences from single-mediator interventional analyses. Indirect effects quantify the portion of the association carried through the specified post-randomisation external referral variable. Negative direct or total effects indicate fewer offending events among placebo participants.

**Supplementary Table 5. Interventional single-mediator analyses of post-randomisation external referrals for violent offending at 24 months**

| Mediator | Method | Indirect effect (95% CI) | Direct effect (95% CI) | Total effect (95% CI) |
|---|---|---|---|---|
| Court assistance/support | Common support | 0.02 [-0.01, 0.06] | -0.23 [-0.47, -0.03] | -0.20 [-0.44, -0.01] |
| Court assistance/support | Entropy balanced | 0.02 [-0.02, 0.06] | -0.28 [-0.59, -0.03] | -0.27 [-0.58, -0.00] |
| Medical services (non-mental health) | Common support | 0.00 [-0.02, 0.03] | -0.20 [-0.45, -0.01] | -0.20 [-0.45, -0.01] |
| Medical services (non-mental health) | Entropy balanced | 0.00 [-0.02, 0.03] | -0.26 [-0.59, -0.00] | -0.26 [-0.60, -0.00] |
| Mental health services | Common support | 0.01 [-0.01, 0.03] | -0.20 [-0.44, -0.01] | -0.20 [-0.44, -0.01] |
| Mental health services | Entropy balanced | 0.01 [-0.01, 0.04] | -0.27 [-0.59, -0.01] | -0.26 [-0.59, 0.00] |
| Legal services | Common support | 0.02 [-0.00, 0.08] | -0.23 [-0.45, -0.04] | -0.21 [-0.44, -0.02] |
| Legal services | Entropy balanced | 0.02 [-0.00, 0.07] | -0.28 [-0.62, -0.04] | -0.27 [-0.60, -0.01] |
| Event count | Common support | 0.04 [-0.00, 0.08] | -0.24 [-0.48, -0.05] | -0.21 [-0.45, -0.01] |
| Event count | Entropy balanced | 0.04 [-0.01, 0.08] | -0.31 [-0.62, -0.06] | -0.27 [-0.60, -0.01] |
| Distinct service categories | Common support | 0.03 [-0.02, 0.08] | -0.23 [-0.47, -0.04] | -0.20 [-0.45, -0.01] |
| Distinct service categories | Entropy balanced | 0.03 [-0.02, 0.08] | -0.29 [-0.60, -0.05] | -0.26 [-0.58, 0.00] |

Estimates are standardised mean count differences from single-mediator interventional analyses. Indirect effects quantify the portion of the association carried through the specified post-randomisation external referral variable. Negative direct or total effects indicate fewer offending events among placebo participants.

**Supplementary Table 6. Interventional single-mediator analyses of post-randomisation external referrals for domestic-violence offending at 12 months**

| Mediator | Method | Indirect effect (95% CI) | Direct effect (95% CI) | Total effect (95% CI) |
|---|---|---|---|---|
| Court assistance/support | Common support | -0.00 [-0.04, 0.04] | -0.41 [-0.68, -0.13] | -0.41 [-0.70, -0.12] |
| Court assistance/support | Entropy balanced | -0.00 [-0.03, 0.04] | -0.37 [-0.71, -0.12] | -0.37 [-0.73, -0.13] |
| Medical services (non-mental health) | Common support | 0.00 [-0.02, 0.02] | -0.42 [-0.71, -0.12] | -0.41 [-0.70, -0.12] |
| Medical services (non-mental health) | Entropy balanced | -0.00 [-0.02, 0.02] | -0.37 [-0.72, -0.12] | -0.37 [-0.75, -0.13] |
| Mental health services | Common support | 0.01 [-0.00, 0.03] | -0.42 [-0.71, -0.12] | -0.41 [-0.71, -0.11] |
| Mental health services | Entropy balanced | 0.01 [-0.01, 0.04] | -0.38 [-0.74, -0.12] | -0.37 [-0.73, -0.11] |
| Legal services | Common support | 0.02 [-0.00, 0.06] | -0.45 [-0.75, -0.14] | -0.43 [-0.71, -0.13] |
| Legal services | Entropy balanced | 0.02 [-0.00, 0.06] | -0.40 [-0.76, -0.13] | -0.38 [-0.74, -0.13] |
| Event count | Common support | 0.03 [-0.01, 0.07] | -0.45 [-0.74, -0.17] | -0.42 [-0.69, -0.13] |
| Event count | Entropy balanced | 0.03 [-0.00, 0.07] | -0.42 [-0.76, -0.16] | -0.38 [-0.74, -0.13] |
| Distinct service categories | Common support | 0.02 [-0.02, 0.07] | -0.43 [-0.71, -0.15] | -0.41 [-0.70, -0.11] |
| Distinct service categories | Entropy balanced | 0.03 [-0.02, 0.07] | -0.40 [-0.72, -0.14] | -0.37 [-0.74, -0.11] |

Estimates are standardised mean count differences from single-mediator interventional analyses. Indirect effects quantify the portion of the association carried through the specified post-randomisation external referral variable. Negative direct or total effects indicate fewer offending events among placebo participants.

**Supplementary Table 7. Interventional single-mediator analyses of post-randomisation external referrals for domestic-violence offending at 24 months**

| Mediator | Method | Indirect effect (95% CI) | Direct effect (95% CI) | Total effect (95% CI) |
|---|---|---|---|---|
| Court assistance/support | Common support | 0.01 [-0.04, 0.07] | -0.50 [-0.81, -0.21] | -0.49 [-0.81, -0.20] |
| Court assistance/support | Entropy balanced | 0.01 [-0.03, 0.06] | -0.46 [-0.84, -0.15] | -0.45 [-0.85, -0.14] |
| Medical services (non-mental health) | Common support | 0.00 [-0.03, 0.03] | -0.49 [-0.80, -0.19] | -0.49 [-0.81, -0.19] |
| Medical services (non-mental health) | Entropy balanced | 0.01 [-0.03, 0.03] | -0.46 [-0.86, -0.15] | -0.45 [-0.85, -0.13] |
| Mental health services | Common support | 0.01 [-0.01, 0.04] | -0.50 [-0.82, -0.20] | -0.49 [-0.81, -0.18] |
| Mental health services | Entropy balanced | 0.02 [-0.01, 0.05] | -0.47 [-0.86, -0.15] | -0.46 [-0.85, -0.13] |
| Legal services | Common support | 0.02 [-0.00, 0.08] | -0.53 [-0.82, -0.21] | -0.50 [-0.81, -0.20] |
| Legal services | Entropy balanced | 0.02 [-0.00, 0.07] | -0.49 [-0.87, -0.17] | -0.47 [-0.85, -0.15] |
| Event count | Common support | 0.04 [-0.01, 0.10] | -0.53 [-0.82, -0.22] | -0.49 [-0.81, -0.19] |
| Event count | Entropy balanced | 0.05 [-0.01, 0.10] | -0.52 [-0.88, -0.20] | -0.47 [-0.85, -0.15] |
| Distinct service categories | Common support | 0.04 [-0.02, 0.09] | -0.52 [-0.81, -0.21] | -0.49 [-0.81, -0.17] |
| Distinct service categories | Entropy balanced | 0.04 [-0.01, 0.10] | -0.50 [-0.87, -0.20] | -0.46 [-0.86, -0.14] |

Estimates are standardised mean count differences from single-mediator interventional analyses. Indirect effects quantify the portion of the association carried through the specified post-randomisation external referral variable. Negative direct or total effects indicate fewer offending events among placebo participants.
